# Methods and computational techniques for predicting adherence to treatment: a scoping review

**DOI:** 10.1101/2024.06.10.24308540

**Authors:** Beatriz Merino-Barbancho, Ana Cipric, Peña Arroyo, Miguel Rujas, Rodrigo Martín Gómez del Moral Herranz, Torben Barev, Nicholas Ciccone, Giuseppe Fico

**Affiliations:** Universidad Politécnica de Madrid, Life Supporting Technologies Research Group, Madrid, Spain; Novo Nordisk A/S, Vandtårnsvej 108-110, 2860, Søborg, Denmark

## Abstract

**Background:** Treatment non-adherence of patients stands as a major barrier to effectively manage chronic conditions. Treatment adherence can be described as the extent to which a patient’s behavior of taking medications follows the agreed recommendations from the healthcare provider. However, non-adherent behavior is estimated to affect up to 50% of patients with chronic conditions, leading to poorer health outcomes among patients, higher rates of hospitalization, and increased mortality. In fact, 200.000 premature deaths each year in the European Union are related to non-adherence. A promising approach to understand adherence behavior of patients represent artificial intelligence and computational techniques. These techniques can be especially useful in analyzing large amounts of heterogeneous patient data, identifying both inter and intra-relationships between factors and patterns associated with non-adherence.

**Objective:** This study offers a provision of a structured overview of the computational methods and techniques used to build predictive models of treatment adherence of patients.

**Methodology:** A scoping review was conducted, and the following databases were searched to identify relevant publications: PubMed, IEEE and Web of Science. The screening of publications consisted of two steps. First, the hits obtained from the search were independently screened and selected using an open-source machine learning (ML)-aided pipeline applying active learning: ASReview, Active learning for Systematic Reviews. Publications selected for further review were those highly prioritized by ASReview.

**Results:** 45 papers were selected into the second round of screening were reviewers performed the full-text screening. The final review included 29 papers. The findings suggest supervised learning (regression and classification) to be the most used analytical approach. Over 54% of adherence topics being related to chronic metabolic conditions such as diabetes, hypertension, and hyperlipidemia. Most assessed predictors were both treatment and socio-demographic and economic-related factors followed by condition-related factors. The selection of a particular computational technique was based on the research question, the type of data available and the desired outcome. A limitation of the reviewed studies is the lack of accountancy for interrelationships between different determinants of adherence behavior. Adherence behavior is a complex phenomenon that is influenced by multiple factors, and it is likely that these factors interact with one another in complex ways.

**Conclusion:** The creation of systems to accurately predict treatment adherence can pave the way for improved therapeutic outcomes, reduced healthcare costs and enabling personalized treatment plans. This paper can support to understand the efforts made in the field of modeling adherence-related factors. In particular, the results provide a structured overview of the computational methods and techniques used to build predictive models of treatment adherence of patients in order to guide future advancements in healthcare.

## Background

The extent to which patients adhere to medical recommendations is a critical factor for successful disease prevention, management and intervention across a wide range of diseases and conditions (Vrijens et al., 2012). However, suboptimal adherence to recommendations is estimated to affect up to 50% of patients with chronic conditions, which results in 125 billion EUR in avoidable premature deaths, hospitalizations and emergency care visits in both EU and US yearly (Lehmann et al., 2014). Addressing the issues of non-adherence would significantly improve individual patient outcomes and reduce related costs. Although several solutions for non-adherence have been developed, evidence of the effects of these solutions is highly variable (Vervloet et al., 2012; Viswanathan et al., 2012), which might be an artifact of the paucity of generalizable knowledge on determinants of adherence behavior in general. In turn, knowledge about determinants of non-adherent behavior could enable healthcare strategies to target these factors in particular, before preventive or treatment measures failure occurs (Haynes et al., 2002).

The multifaceted nature of adherence behavior made an investigation into determinants of adherence behavior a complex task. There has been a vast of research conducted finding an ambiguous impact of a number of different factors. The ambiguity in findings could be due to a lack of accountancy for interrelationships between different determinants of adherence behavior. Artificial intelligence (AI) bears the potential to provide an understanding of adherence behavior by analyzing large amounts of heterogeneous patient data in identifying both inter and intra-relationships between factors and patterns associated with non-adherence. To understand the efforts made in the field of modeling adherence-related factors and in order to guide future advancements, the aim of this article is to provide an overview of computational techniques which have been employed so far.

### Study objective and research question

The main objective of this article is to conduct a scoping review to identify the computational techniques and their features that are implemented in treatment adherence models. Specifically, the objectives were to review the literature to identify features and evidence-based results that have demonstrated relevance to the implementation of these automated models for this purpose. Ultimately, the idea behind these scoping reviews was to have a structured overview of the key concepts in the research area, identify gaps in the existing research and succinctly summaries the key findings. This scoping review is based in the following research question:

- What are the computational methods and techniques used to build predictive models of treatment adherence?

## Materials and methods

### Scoping approach: sources and keywords

To conduct the search on computational methods and techniques employed in predictive modeling of treatment adherence, the preselected information sources were electronic databases of scientific publications, PubMed, IEEE and Web of Science in particular.

A series of keywords were selected and grouped into three groups:

- Theoretical models: Covering current theories and models on adherence behavior (e.g., “transtheoretical model”, “ABC Taxonomy”).
- Computational techniques: Covering the most relevant computational techniques related to artificial intelligence (e.g., “artificial intelligence”, “machine learning”, “classification”).
- Adherence field: Encompassing the most commonly used terms that represent the term adherence to treatment (e.g., “treatment adherence”, “patient compliance”).

### Search strategy

In order to identify relevant publications on computational methods/techniques for predicting adherence to treatment we performed a search across the three databases mentioned in the previous section. The search queries were adapted to each database to ensure comprehensive and right extraction of the literature, using both MeSH Terms and free-text keywords for PubMed, full-text and metadata searches for IEEE Xplore and topic searches for Web of Science. Table 1 includes the specific search queries used for each dataset, categorized into theoretical models, computational techniques and adherence-related terms.

**Table 1.**
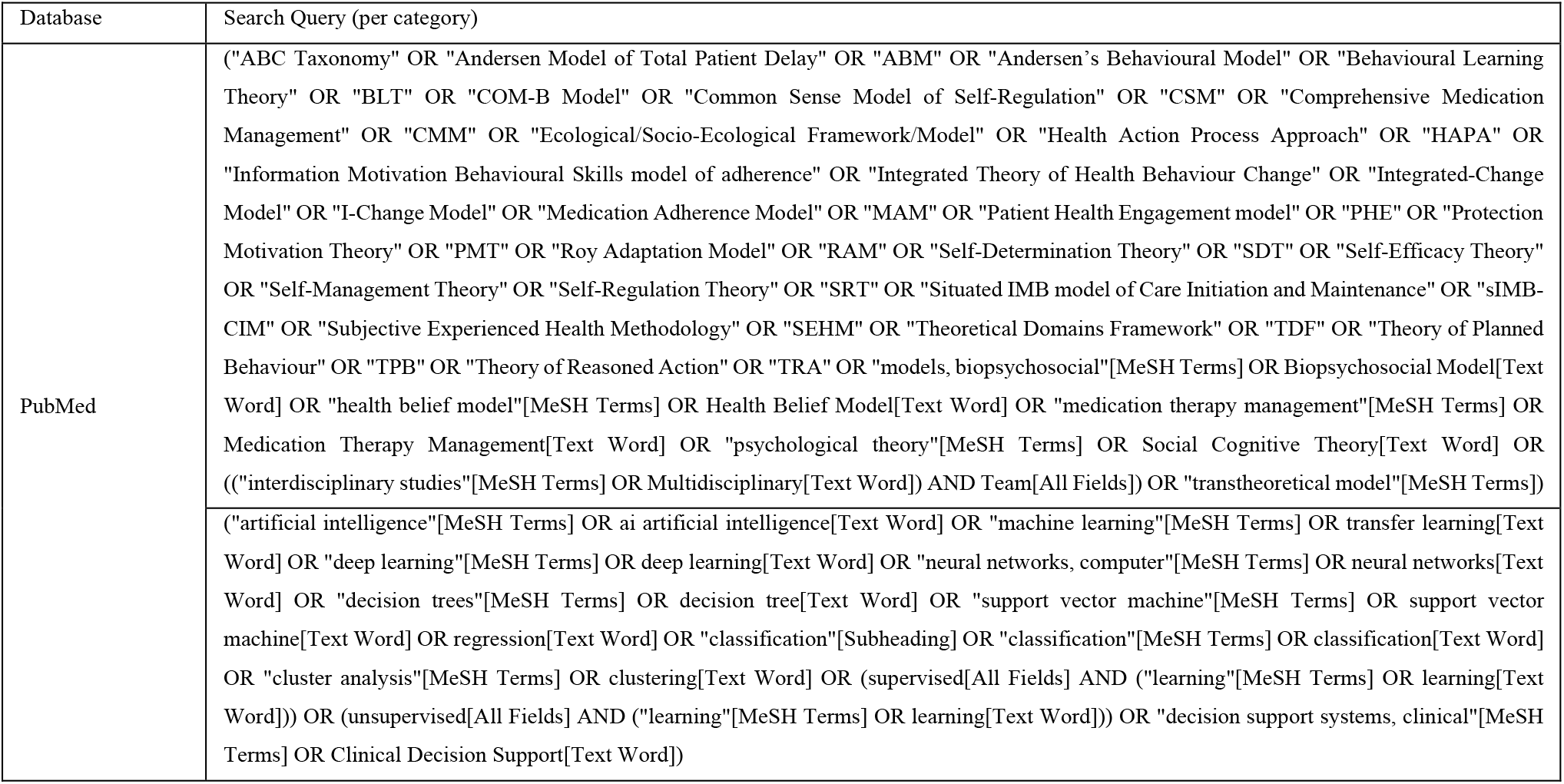
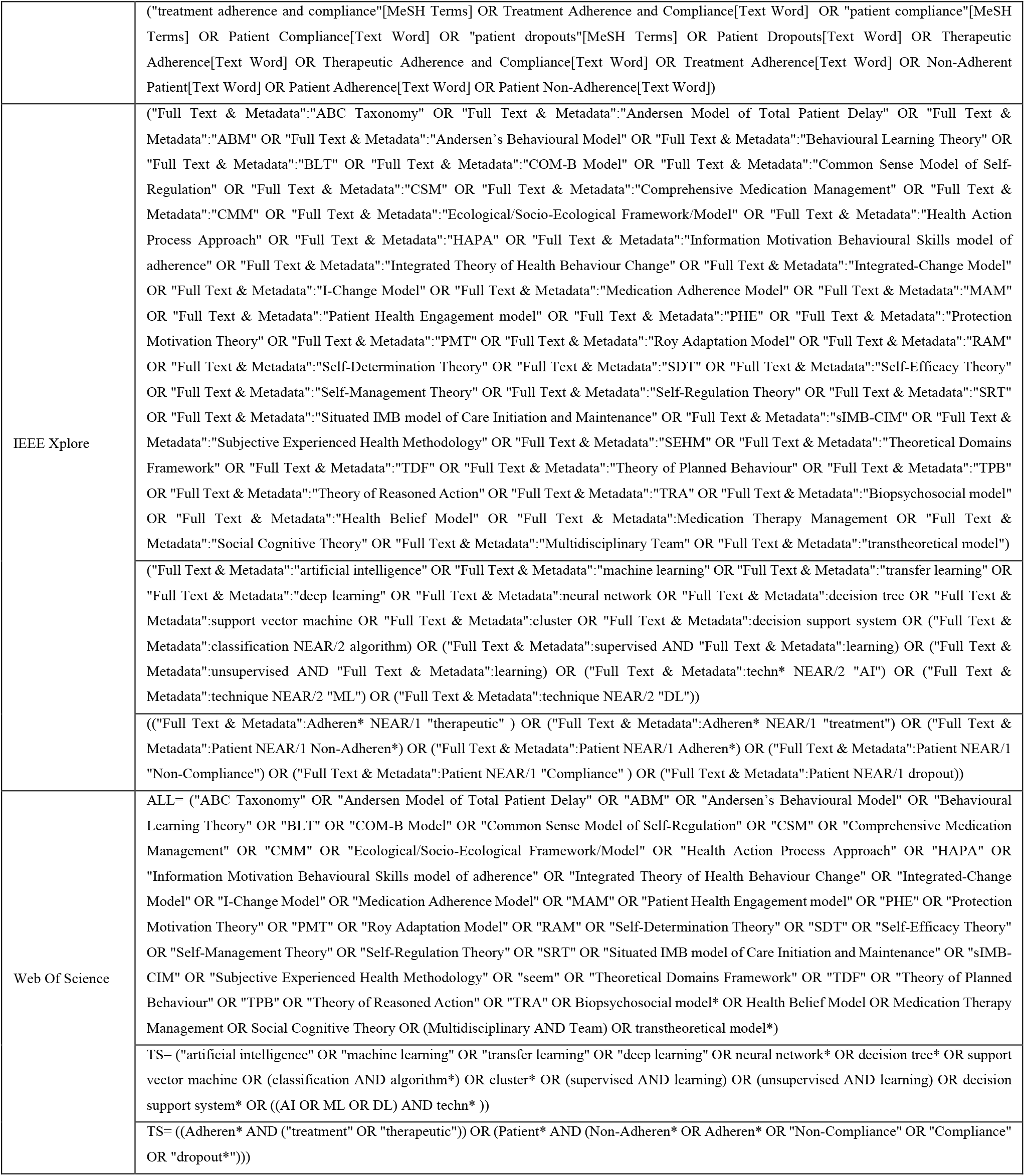
Databases search queries.

### Eligibility criteria

The eligibility criteria have been defined using JBI methods for a scoping review (Peters et al.,2020) in order to guide the selection process. The criteria were divided into the following areas:

- Population: Studies considering adult human subjects (≥ 16 years old).
- Concept: Computational methods and techniques for treatment adherence prediction models.
- Timeframe: Within the last 11 years (2012-2023).
- Language: Articles only published in English.

### Data extraction

The data was extracted using Microsoft Excel. The search hits (including publication title, authors, abstract and DOI) are downloaded in .csv, .txt or .xlsx format, merged and cleared of duplicates. The title and abstract screening were performed using ASReview software (*ASReview: AI-Aided Open Source Systematic Review Software*, n.d.)and stored in the excel file for the text review. Reviewers, including biomedical engineers, behavioral data scientists, psychologists, pharmacologists and doctors, manually annotated the following publication details: the type of study/model and publication, the objective of the study, the type and the data used, the theoretical model behind, the disease that each article focuses, the type of methodology/technique used, the results obtained, the limitations of the study and proposed future research.

### First screening: title & abstract

The hits obtained from the search will be independently screened and selected using ASReview. The tool is initially trained with 10 relevant and 10 irrelevant publications selected by two independent researchers. After feeding the tool with the training publications, the tool returns the set of hits ordered according to relevance priority. These results are checked by an independent researcher. In case of several irrelevant results among the top priority hits, the tool is further trained by manually screening at least 1% of the total number of publications in the whole set. Publications selected for further full-text review were those highly prioritized by ASReview. After this, each resulting abstract is consulted by two independent reviewers who must decide whether or not to include the article for the full text review. Discrepancies between them were analyzed by a third independent reviewer who acted as a referee.

### Second screening: full text

Each reviewer performed the full-text screening of the assigned publications considering the eligibility criteria described above. For each publication assigned, the reviewer checked each eligibility criteria and consider for further inclusion only those publications meeting all the criteria. For the selected publications, reviewers will annotate additional publication details as requested on the spreadsheet. After the full-text screening, the final list of selected publications was ready for data extraction.

### Outcomes

The final list of included studies for the ScR (Scoping Review) was divided among several experts who worked in parallel to extract the relevant data from the assigned publications. To facilitate and homogenize the process of data extraction, a structured data collection sheet was shared among reviewers, in which the elements considered of interest in this scoping review were included. The articles for which data was extracted were articles that mentioned the architecture and computational techniques employed in the development of treatment adherence prediction models. Factors extracted from these articles were: medical condition, a measure of adherence as an outcome variable, sample size, adherence-related factors (e.g., socio-demographic, healthcare system-related, condition-related, treatment-related, and patient-related), computational technique and computational algorithm. Socio-demographic and economic factors refer to characteristics such as age, gender, income and education level. Healthcare system-related factors refer to descriptors of the healthcare environment, such as HC visits and interactions. Condition-related factors refer to aspects of the patient’s health condition, such as symptom severity, which may impact adherence. Treatment-related factors refer to aspects of the treatment regimen such as complexity, dosing schedule, and side effects, which may impact adherence. Patient-related concern inter and intra-personal characteristics of the patient.

Computational algorithms used were classified as clustering, classification and regression, depending on whether they were aimed at detecting association, classification or clusterisation.

### Materials

This study utilized a multifaceted approach integrating various technological tools. First, we employed the BEAMER project (*_BEAMER HOME - BEAMER*, n.d.), funded through the Innovative Medicines Initiative (IMI2) and now under the Innovative Health Initiative (IHI), is a European research project (Grant agreement number: 101034369) dedicated to developing a behavioral and adherence model leveraging artificial intelligence (AI) techniques. BEAMER’s primary aim is to enhance the quality, outcomes, and cost-effectiveness of healthcare services. Secondly, we incorporated the Active Learning for Systematic Reviews (ASReview) tool. ASReview is an open-source machine learning (ML)-aided pipeline that applies active learning methodologies. It significantly improves the efficiency of screening academic titles and abstracts by strategically prioritizing them through active learning algorithms. Lastly, Microsoft Excel, a widely recognized software program created by Microsoft, was utilized. Excel’s powerful spreadsheet capabilities, which include organizing numbers and data with formulas and functions, were crucial in data analysis and management phases of the research.

## Results

Of 45 papers selected into the second round of screening, the final review included 29 papers (Figure 1)(Table 2). A total of twelve papers were excluded due to being randomized control trials (9), employing qualitative methodology (4), being a literature review of adherence factors without reference to analytical techniques (1) and/or for having different outcome variables in focus (2; Figure 1). With respect to analytical techniques, three major approaches were identified; generalized linear models accounted for 21,67% of employed techniques, followed by the family of logistic regressions (20,00%) and random forest techniques (18,33%; Table 3). The findings suggest supervised learning (regression and classification) to be the most used analytical approaches. The family of generalized linear models employed included multiple, multilevel, hierarchical, fixed-effects, OLS, mixed-effects and GEE (Table 3 and Table 4). Whether the selected algorithm was a regression or classification, was primarily determined by the data source and the scaling of the outcome and predictors measures (Table 2). The same applied to a selection of logistic regression or random forest techniques.

**Table 2.** Description of Articles *.

| Authors, Year | Condition | Adherence (Outcome Variable) | Sample Size | Main Computational Technique | Computational Algorithm | ADHERENCE FACTORS: Socio-demographic | Healthcare System | Condition-related | Treatment-related | Patient-related |
| --- | --- | --- | --- | --- | --- | --- | --- | --- | --- | --- |
| (Shiyanbola et al., 2018) | Type 2 Diabetes | Categorical (Self-reported) / The Morisky Medication Adherence Scale (MMAS-8) | 174 | Cluster Analysis | Clustering | + |  | + | + | + |
| (Colvin et al., 2018) | Diabetes, Hypertension, or Hyperlipidemia | Continuous / Pharmacy claims data - number of medication days | 56 | Linear Regression | Regression | + | + | + | + | + |
| (Fortuna et al., 2018) | Hypertension | Categorical (Self-reported) / The Morisky Medication Adherence Scale (MMAS-8) | 2128 | Logistic Regression | Classification | + |  |  | + |  |
| (Wang et al., 2020) | Crohn's Disease | Categorical (Self-reported) / The Medication Adherence Report Scale (MARS) | 446 | Random Forest, Back-propagation Neural Network, Support Vector Machine, Logistic Regression | Classification, Regression | + | + | + | + | + |
| (Wawruch et al., 2019) | Peripheral Arterial Disease | Dichotomous (Adherent/Non-adherent) / 6-month period without statin prescription | 8330 | Cox Regression | Regression | + |  | + | + |  |
| (Whittle et al., 2016) | Hypertension, Hyperlipidemia | Dichotomous (Adherent/Non-adherent) / <80 % of medication refilled | 31250 | Logistic Regression, Linear Regression (multivariate) | Classification, Regression | + |  |  |  |  |
| (Wong et al., 2012) | Diabetes | Dichotomous (Adherent/Non-adherent) / <80 % of medication refilled | 444418 | Logistic Regression (hierarchical, multilevel) | Classification | + | + | + | + | + |
| (Wu et al., 2020) | Type 2 Diabetes | Dichotomous (Adherent/Non-adherent) / <80 % of medication refilled | 401 | Logistic Regression, C 5.0 model, Decision List, Bayesian Network, Discriminant model, KNN algorithm, LSVM, Random Forest, SVM, Tree-AS, CHAID, Quest, C&R Tree, Neural Net, and the Ensemble model | Classification | + |  | + | + | + |
| (Koulayev et al., 2017) | Hypertension, Diabetes, Dyslipidemia | Continuous / Number of refills ('medication possession ratio' (MPR)) | 15916 | OLS Regression (fixed-effects) | Regression | + | + | + | + |  |
| (Kozma et al., 2014) | Multiple sclerosis | Continuous / Number of refills | 4606 | Linear Regression (multiple) | Regression | + |  | + | + |  |
| (Alatawi et al., 2016) | Type 2 Diabetes | Categorical (Self-reported) / three different adherence measures | 222 | Linear Regression (multiple) | Regression | + | + | + | + | + |
| (Berry et al., 2015) | Cancer | Categorical Dichotomized (Self-reported) / The Morisky Medication Adherence Scale (MMAS-8) - categorized | 752 | Logistic Regression (univariate), Classification Tree | Classification | + | + | + | + |  |
| (Boland et al., 2016) | Chronic Obstructive Pulmonary Disease | Dichotomous (Adherent/Non-adherent) / <80 % of medication refilled | 511 | Linear Regression (mixed-effect) | Regression | + |  | + | + | + |
| (Campbell et al., 2020) | Hypertension | Dichotomous (Adherent/Non-adherent) / <80 % of medication refilled | 4842 | Generalized Linear Models (with log link, gamma or negative binomial distribution) | Regression | + | + | + | + |  |
| (Horberg et al., 2012) | HIV-positive | Categorical / prescription based treatment regimens | 10801 | Regression Tree, Linear Regression (mixed-effect), Logistic Regression (mixed-effect) | Regression, Classification | + | + | + | + |  |
| (Javanmard ifard et al., 2020) | Type 2 Diabetes | Categorical (Self-reported) / The Morisky Medication Adherence Scale (MMAS-8) | 227 | Correlations | Description | + | + |  | + |  |
| (Sleath et al., 2015) | Glaucoma | Continuous (Self-reported) | 279 | Generalizing Estimating Equations | Regression | + | + |  | + | + |
| (Tibbitt et al., 2020) | Asthma | Continuous / Doses taken (Percentage) | 220 | Principal Component Analysis, K-Means, Decision Trees | Classification, Regression, Clustering |  |  | + | + |  |
| (Tiv et al., 2012) | Type 2 Diabetes | Categorical (Self-reported) / Six-items questionnaire - 'good', 'medium' and 'poor' (treated as nominal) | 3637 | Logistic Regression (multinomial polychotomous) | Classification | + | + | + | + |  |
| (Verma et al., 2018) | Hypertension | Dichotomous (Adherent/Non-adherent Medication discontinuation) | 13350 | Cox proportional hazards regression (propensity score matched sample) | Classification, Regression |  |  |  | + |  |
| (Hsu et al., 2022) | Cardiovascular disease | Dichotomous, Adherent/Non-adherent // Proportion of days covered (PDC) $\geq 80$ is used as the threshold for adherence | 100096 | LSTM, Simple RNN, MLP, Ridge Classifier, Logistic Regression | Regression, Classification | + | + | + | + | |
| (Senner et al., 2023) | Schizophrenia -spectrum and bipolar disorder | Categorical / The medication adherence was evaluated with a self-rating questionnaire with assemblance to BARS | 862 | Linear Regression (multiple), Cluster analysis (hierarchical) | Regression, Clustering | + |  | + | + |  |
| (Innab et al., 2023) | Hypertension | Continuous / Correlations between adherence and some factors | 114 | Pearson correlation, Linear regression (hierarchical) | Regression | + |  | + | + | + |
| (Luo et al., 2023) | Breast Cancer | Dichotomous, Poor/Good functional exercise compliance // Based on the program of their hospitals | 227 | Classification Tree (CHAID) | Classification | + |  | + |  | + |
| (Carmody et al., 2023) | Sleep disordered breathing (SDB) | Continuous / Relationships between PAP frequency/duration and some factors | 188 | Linear Regression (multiple) | Regression | + |  | + | + | + |
| (Hu et al., 2022) | Breast Cancer | Dichotomous, Adherent/Non- adherent / Adherence as the proportion of days covered (PDC) by AET | 559 | Linear Regression (multivariate) | Regression | + |  | + | + | + |
| (Malo et al., 2022) | Cardiovascular disease | Continuous / Correlations between adherence and some factors | 15332 | Logistic Regression (multinomial) | Regression | + |  | + | + |  |
| (Li et al., 2022) | Type 2 Diabetics | Dichotomous, Adherent/Non-adherent / Proportion of days covered (PDC) $\geq 80$ is used as the threshold for adherence | 980 | ANOVA | Regression | + | | + | + | + |
| (Browne et al., 2023) | Prevention of Immunodeficiency Virus Infection | Dichotomous, Adherent/Non-adherent, Continuous / Taking and timing | 63 | Logistic Regression (mixed-effects) | Regression | + |  | + | + | + |

\* Author, Year: Authors and year of publication for the study, Condition: disease under investigation, Adherence (outcome variable): feature measuring adherence, Sample Size: size of the sample in the study, Main Computational Technique: primary algorithm used in the analysis, ADHERENCE FACTORS: factors considered in the study, categorized into socio-demographic, healthcare system, condition-related, treatment-related, and patient-related, + means the presence of associated factors.

**Table 3.**
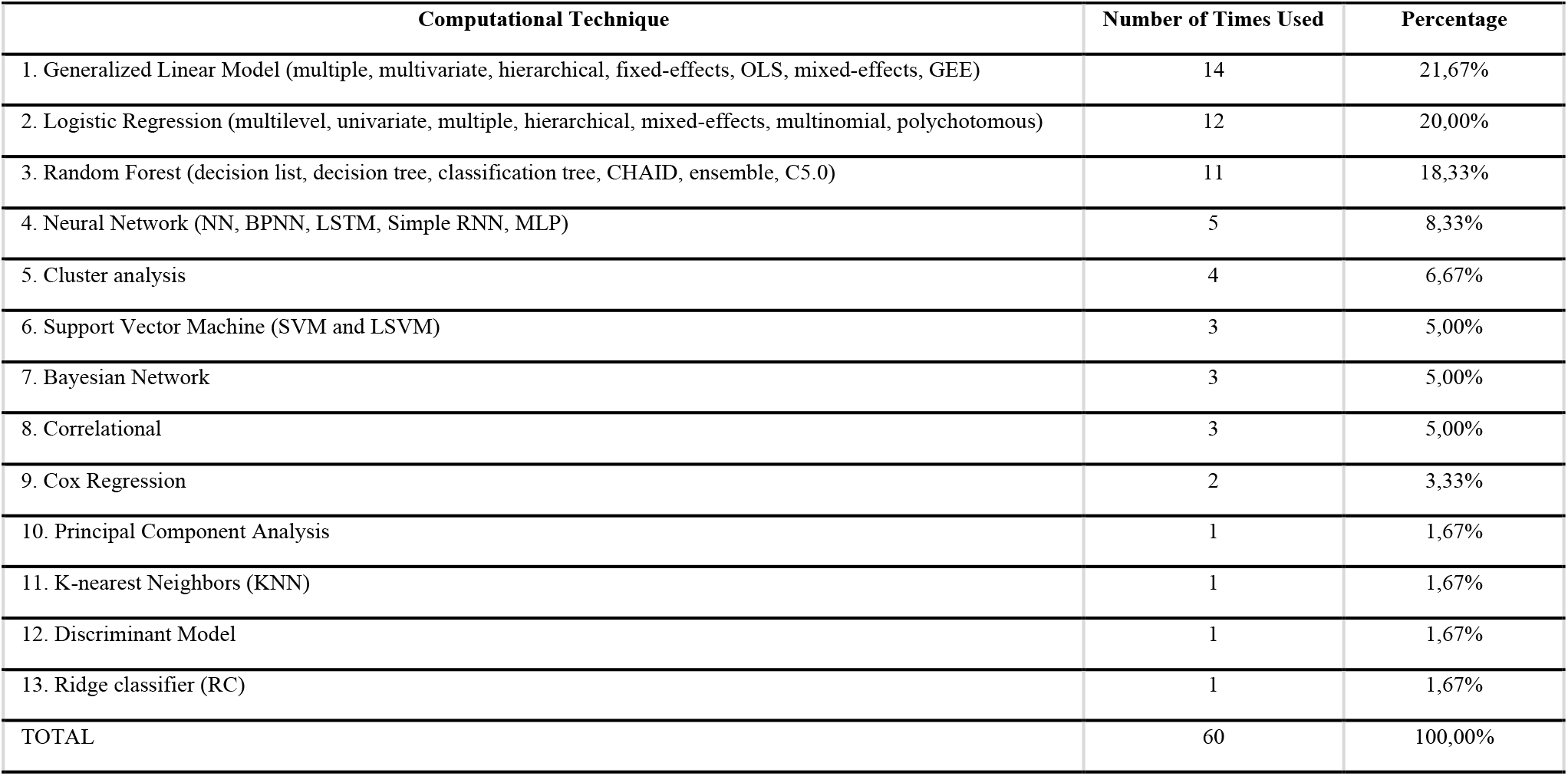
Description of the Computational Techniques.

| Computational Technique | Number of Times Used | Percentage |
| --- | --- | --- |
| 1. Generalized Linear Model (multiple, multivariate, hierarchical, fixed-effects, OLS, mixed-effects, GEE) | 14 | 21,67% |
| 2. Logistic Regression (multilevel, univariate, multiple, hierarchical, mixed-effects, multinomial, polychotomous) | 12 | 20,00% |
| 3. Random Forest (decision list, decision tree, classification tree, CHAID, ensemble, C5.0) | 11 | 18,33% |
| 4. Neural Network (NN, BPNN, LSTM, Simple RNN, MLP) | 5 | 8,33% |
| 5. Cluster analysis | 4 | 6,67% |
| 6. Support Vector Machine (SVM and LSVM) | 3 | 5,00% |
| 7. Bayesian Network | 3 | 5,00% |
| 8. Correlational | 3 | 5,00% |
| 9. Cox Regression | 2 | 3,33% |
| 10. Principal Component Analysis | 1 | 1,67% |
| 11. K-nearest Neighbors (KNN) | 1 | 1,67% |
| 12. Discriminant Model | 1 | 1,67% |
| 13. Ridge classifier (RC) | 1 | 1,67% |
| TOTAL | 60 | 100,00% |

**Table 4.** Adherence Models inspected related to the Conditions.

| Condition | Number of times researched | Percentage of times researched |
| --- | --- | --- |
| Diabetes | 9 | 25,71% |
| Hypertension | 7 | 20,00% |
| Hyperlipidaemia/Dyslipidaemia | 3 | 8,57% |
| HIV + | 2 | 5,71% |
| Breast Cancer | 2 | 5,71% |
| Crohn's disease | 1 | 2,86% |
| Peripheral arterial disease (PAD) | 1 | 2,86% |
| Chronic Obstructive Pulmonary Disease (COPD) | 1 | 2,86% |
| Cancer (general) | 1 | 2,86% |
| Asthma | 1 | 2,86% |
| Glaucoma | 1 | 2,86% |
| Multiple sclerosis | 1 | 2,86% |
| Schizophrenia-spectrum disorder | 1 | 2,86% |
| Bipolar disorder | 1 | 2,86% |
| Primary cardiovascular disease | 1 | 2,86% |
| Cardiovascular disease (general) | 1 | 2,86% |
| Sleep disordered breathing (SBD) | 1 | 2,86% |
| Total | 35 | 100,00% |

**Figure 1.**
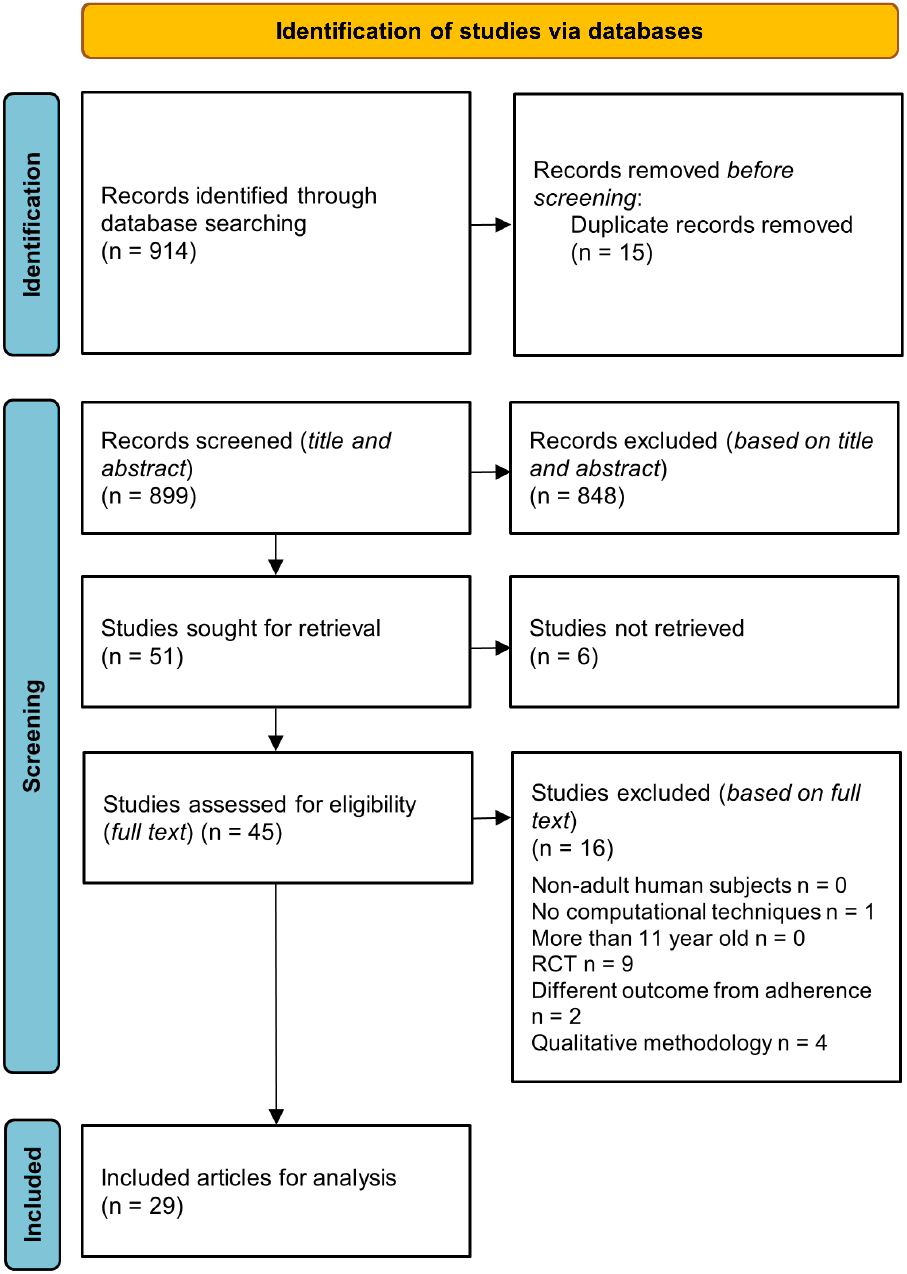
Flow Diagram

Table 4 shows a distribution of treatments of interest in relation to adherence, with over 54% of adherence topics being related to chronic metabolic conditions such as diabetes, hypertension, and hyperlipidemia. The other conditions and equally distributed among each other, were HIV, Breast Cancer, Peripheral Arterial Disease (PAD), Chronic Obstructive Pulmonary Disease (COPD), Cancer (general), Asthma, Glaucoma, Multiple sclerosis, some mental disorders, Crohn’s disease, some other cardiovascular diseases and Sleep disordered breathing (SDB). Most assessed predictors were both treatment and socio-demographic and economic-related factors, followed by condition-related factors. Somewhat less inspected are factors related to the healthcare system and patient-related individual differences (see Table 1). The outcome adherence to the treatment variable was uniformly specified as dichotomous (12), categorical (9) and continuous (8) of the analyzed articles (Table 1). An example of categorical indicator for adherence was mostly The Morisky Medication Adherence Scale (MMAS-8), a self-reported measure of adherence. Some continuous and dichotomous indicators were based on the prescriptions regiments and the number of pharmacy claims. Dichotomous indicators classified adherence when a proportion of days covered by prescription/pharmacy claims equalled or exceeded 80%. MPR (medical possession ratio and PDC (proportion of days covered by prescription) are almost identical and equally common formula-based calculations of adherence (and the 80% threshold).

## Discussion

Based on the results of the review, to this date different computational techniques have been employed to model treatment adherence. The review included 29 papers of which over three quarters employed generalized linear models, logistic regressions and random forest techniques. Supervised learning (regression and classification) was the most used analytical approach. The review also revealed that chronic metabolic conditions such as diabetes, hypertension and hyperlipidaemia were the most common conditions for which adherence was modelled. The predictors assessed in the studies were treatment-related factors, socio-demographic and economic factors and condition-related factors, with healthcare system and patient-related individual differences being somewhat less inspected.

The selection of a particular computational technique was based on the research question, the type of data available and the desired outcome. For example, regression techniques such as linear regression or logistic regression were commonly used when the outcome variable was continuous or dichotomous, respectively. Random forest techniques, on the other hand, were used when dealing with large, complex datasets that may contain many predictor variables. Indeed, the predictors assessed in the studies included several different factors.

However, one limitation of the reviewed studies is the lack of accountancy for interrelationships between different determinants of adherence behavior. Adherence behavior is a complex phenomenon that is influenced by multiple factors and it is likely that these factors interact with one another in complex ways. For example, a patient’s socio-economic status may affect their ability to afford medication, which in turn may impact their adherence. Additionally, a patient’s health condition may impact their ability to adhere to certain aspects of the treatment regimen, such as exercise or dietary restrictions. To address this limitation, future research could consider more comprehensive models that consider the interrelationships between different determinants of adherence behavior. This could involve the use of more advanced analytical techniques such as network analysis or systems modelling. Additionally, researchers could explore the use of machine learning algorithms that are better able to capture complex interactions between multiple predictors. This could ultimately lead to more effective interventions that are better tailored to individual patients’ needs and circumstances. The results suggest that computational techniques have been useful in modelling treatment adherence for a variety of conditions. However, the review also highlights the need for further research to address the limitations and challenges in the current approaches and to refine and validate computational models of treatment adherence.

## Conclusion

Treatment adherence behaviour is a complex phenomenon that is influenced by multiple factors. This is critical, as treatment non-adherence stands as a major barrier to effectively manage chronic conditions. A promising approach to understand adherence behaviour of patients in detail and analyse large amounts of heterogeneous patient data represent artificial intelligence and computational techniques. These techniques can be especially fruitful in identifying both inter and intra-relationships between factors and patterns associated with non-adherence.

This article sheds light of the computational methods and techniques used to build predictive models of treatment adherence. In particular, the results of the conducted scoping review provide a structured overview of the key concepts in the research area is provided, gaps in the existing research were identified and the key findings succinctly summarised.

These results of this paper contribute to understand the efforts made in the field of modelling treatment adherence-related factors and to guide future advancements. An advancement of knowledge of treatment adherence behaviour can make healthcare more efficient, can contribute to deliver high-value personalised care, increase the treatment adherence of patients, and result in a significant decrease in healthcare costs.

### Limitations and Future Research

To cover the dominant literature of peer-reviewed published papers in the selected field, the scoping review was limited to three electronic databases of scientific publications: PubMed, IEEE, and Web of Science. Only articles in English language have been included in the analysis. It must be noted that further studies can investigate additional data databases, data sources and articles published in different languages for their analysis. For structuring the results, different grouping and ranking strategies might have also added additional insights but were not attempted since they are outside the scope of this paper.

## Data Availability

All data produced in the present work are contained in the manuscript

## Acknowledgment

The project has received funding from the Innovative Medicines Initiative 2 Joint Un-dertaking under grant agreement No 101034369. This joint undertaking receives sup-port from the European Union’s Horizon 2020 Research and Innovation Pro-gramme, the European Federation of Pharmaceutical Industries and Associations (EFPIA) and Link2Trials. This communication reflects the views of the authors and neither the IMI nor the European Union, EFPIA, or Link2Trials are liable for any use that may be made of the information contained herein. We would like to thank the consortium partners who actively participated in the scouting of articles in phases 1 and 2 and made this study possible.

